# Demography and outcomes of frozen tongue: a scoping review of Scandinavian tundra tongue cases

**DOI:** 10.1101/2025.05.07.25326467

**Authors:** Anders Hagen Jarmund, Sofie Eline Tollefsen, B. Cristoffer Sakshaug, Yashar Honarmandi, Sverre Helge Torp

## Abstract

Children occasionally adhere their tongues to cold metal surfaces during winter (“tundra tongue”), but little is known about the epidemiology and outcomes of these cases. We therefore conducted a scoping review to explore the following questions: who experience tundra tongue, under which circumstances does it occur, and what are the outcomes? Systematic searches were performed to identify case reports published in historical newspapers from Norway, Sweden and Denmark using national library databases. Epidemiological data were charted manually from newspaper items describing actual (not metaphorical) frozen tongues. Among 17 009 unique search hits, 856 reports of 113 different cases were found. Almost all cases (96%) involved children, median age 5.25 years, and boys were in majority (63%). Tongues were most often frozen to railings (40%). Ambient temperature was reported in 18 cases with median -16.5 °C. Outcomes ranged from discomfort to potential amputation of tongue tissue, with 20 (18%) cases involving a doctor or a hospital. Severe injuries were reported in several cases through three distinct mechanisms: (1) the direct effect of cold on tissue, (2) detachment injuries, and (3) the consequences of immobility. In conclusion, parents, health care professionals and policy makers should not underestimate the potential harm of the tundra tongue.

## Introduction

Children sometimes freeze their tongues to cold metal surfaces in winter. While recognized anecdotally, this phenomenon is seldom studied scientifically; only a single case report exists, which described the author’s son and introduced the term “ tundra tongue” (1). Despite this research gap, concerns over the potential risks of cold metal on playgrounds contributed to Norway implementing regulations requiring insulation on playground equipment in the late 1990s (2). Public debates surrounding the regulation explicitly mentioned the risk of children adhering their tongues to metal (3). Critics argued that the measures were overly protective (4), with some suggesting that such incidents could be valuable learning experiences (5), and that the perceived risk was unsupported by data (6).

This scarcity of case reports mirrors a broader gap in research concerning the human tongue’s response to cold exposure. While some studies investigated cold-induced morphological changes in animal tongues (7,8), direct data on human tongue responses are scarce. Oral frostbite injuries in youth and young adults have been reported following inhalation of propellants (9,10) and lip injuries from prolonged contact with ice cream (11). Research on adhesion to cold surfaces has focused on other tissues; for example, models estimate the frostbite injury on the fingertip occurs within seconds on contact with aluminum below -10 °C (12), and studies using pig skin show adhesion forces of 7-9 N on various metals at -40 °C (13). However, the tongue’s moist, mucous surface is likely to cause strongly and perhaps more readily adhesion to cold surfaces compared to drier tissues such as skin.

To address the knowledge gap regarding the “ tundra tongue,” we conducted structured searches in national databases of Scandinavian newspapers to systematically identify and characterize reported incidents. Using a scoping review approach, we aimed to identify reported cases and explore the following questions: Who experiences tundra tongue, under what circumstances does it occur, and what are the outcomes?

## Methods

This paper is guided by the Preferred Reporting Items for Systematic reviews and Meta-Analyses extension for Scoping Reviews (PRISMA-ScR) checklist (14). The protocol for this review is not published.

### Literature search

We performed structured literature searches in the digital newspaper databases of the national libraries of Norway, Sweden and Denmark. Proximity searches were used to identify relevant items, where two keywords and a maximum word distance between them are defined. For example, a proximity search for “ tongue” and “ stuck” with distance 2 detects both “ the tongue got stuck” and “ the tongue immediately got stuck” . We initially tested selected Norwegian search terms, increasing the proximity distance stepwise from 1 to 10 words, to determine the optimal distance for maximizing relevant hits.

Subsequently, we performed comprehensive searches in Norwegian, Swedish, and Danish using this optimal proximity setting. Table S1 provides a complete list of all search terms, including proximity. Initial searches occurred February 7^th^, 2025, updated March 1^st^, 2025, with no publication year restrictions applied.

Access and full-text availability varied between databases. The National Library of Norway (Nasjonalbiblioteket) offers unlimited online access to digitized newspapers for Norwegian researchers by application (15). The database is described as almost complete, covering newspapers from 1763 and onwards. The public Application Programming Interface (API) was used to conduct the searches and collect relevant metadata (16). The National Library of Sweden (Kungliga biblioteket) offers access to a text fragment surrounding the search words, in addition to full-text of newspapers older than 100 years (17). More recent newspapers can only be accessed in person at selected libraries in Sweden. Text fragments were therefore screened online, and potentially relevant items were evaluated in full text at the Kungliga biblioteket, Stockholm, Sweden. The National Library of Sweden covers 400 years of newspapers but has not yet digitalized all newspapers from the period 1907-2013 (17). Danish newspapers were available through the Royal Danish Library’s (Det Kongelige Bibliotek) Mediestream service (18). The database is described as incomplete (19) but covering newspapers from the 1660s and onwards. The Royal Danish Library only offers access to newspaper older than 100 years, and while the searches also returned more recent hits, these were unavailable for assessment.

### Study selection

We included newspaper items describing an actual (i.e., non-metaphorical) frozen tongue. Items were excluded if they 1) did not involve humans, 2) did not refer to actual and specific incidents (e.g., general warnings), or 3) referred to incidences occurring more than a year before the newspaper item was published. A.H.J. screened all items; borderline cases were resolved by consensus among A.H.J., S.E.T., and B.C.S.

### Data charting and statistics

Two authors (A.H.J. and S.E.T.) manually reviewed the included newspaper items and recorded the following data: age (years, child/adult status), gender, ambient temperature, location, date (month/year), object involved, detachment remedy, and outcome (free text, involvement of doctor/hospital/police/fire service). When intervals were reported for age or temperature, the mean value was used for summary statistics. All figures were created using the ggplot2 package (v. 3.5.1) for R (v. 4.4.3) (20,21).

### Declaration of AI and AI-assisted technologies in the writing process

During manuscript preparation, the authors utilized the AI language model Gemini (2.5 Pro, experimental 03-25) by Google to enhance clarity and readability. Selected paragraphs were drafted by the authors and the text provided to the Gemini service using this prompt: “ Please revise the following paragraphs for clarity and readability, flow and structure; grammar, and adherence to scientific writing standards. Start by assessing overall structure, flow, and transitions. Active voice and short, clear sentences are preferred.

Special attention should be given to the first and last sentence of the paragraphs. Provide critical comments and suggestions for improvements. The paragraph is from the [Introduction, Methods, …] section of a research paper.” Gemini suggested improvements that were critically reviewed by the authors and incorporated in the manuscript when appropriate. After using this tool, the authors reviewed and edited the content as needed and take full responsibility for the content of the publication.

### Ethics

This study analyzed publicly available information from editorially controlled media (historical newspapers). Because these data are considered publicly accessible and freely usable for research (22), formal ethical approval was deemed unnecessary. To protect individual privacy, particularly for children involved, case locations are reported only at the country level.

## Results

### Search results

The number of search hits increased considerably at proximities 1 to 3 (Figure S1) while the proportion of new items being included went from 11% at proximity 1, to 13% at proximity 2, and to 3% at proximity 3. At proximities 4-10, the number of new hits was low (Figure S1), and the proportion of new items being included stayed below 3%. Thus, a proximity of 3 was considered optimal and used for the search in Swedish and Danish newspapers.

Across all databases, the searches generated 81,355 results (Figure 1). The full dataset is available (see Data availability). After removing duplicates and 4,209 unavailable items (primarily recent Danish newspapers), 12,800 unique items were screened for eligibility. This screening yielded 856 newspaper items (7% of screened) for inclusion and data charting, with 37 borderline items resolved by author consensus.

**Fig. 1.**
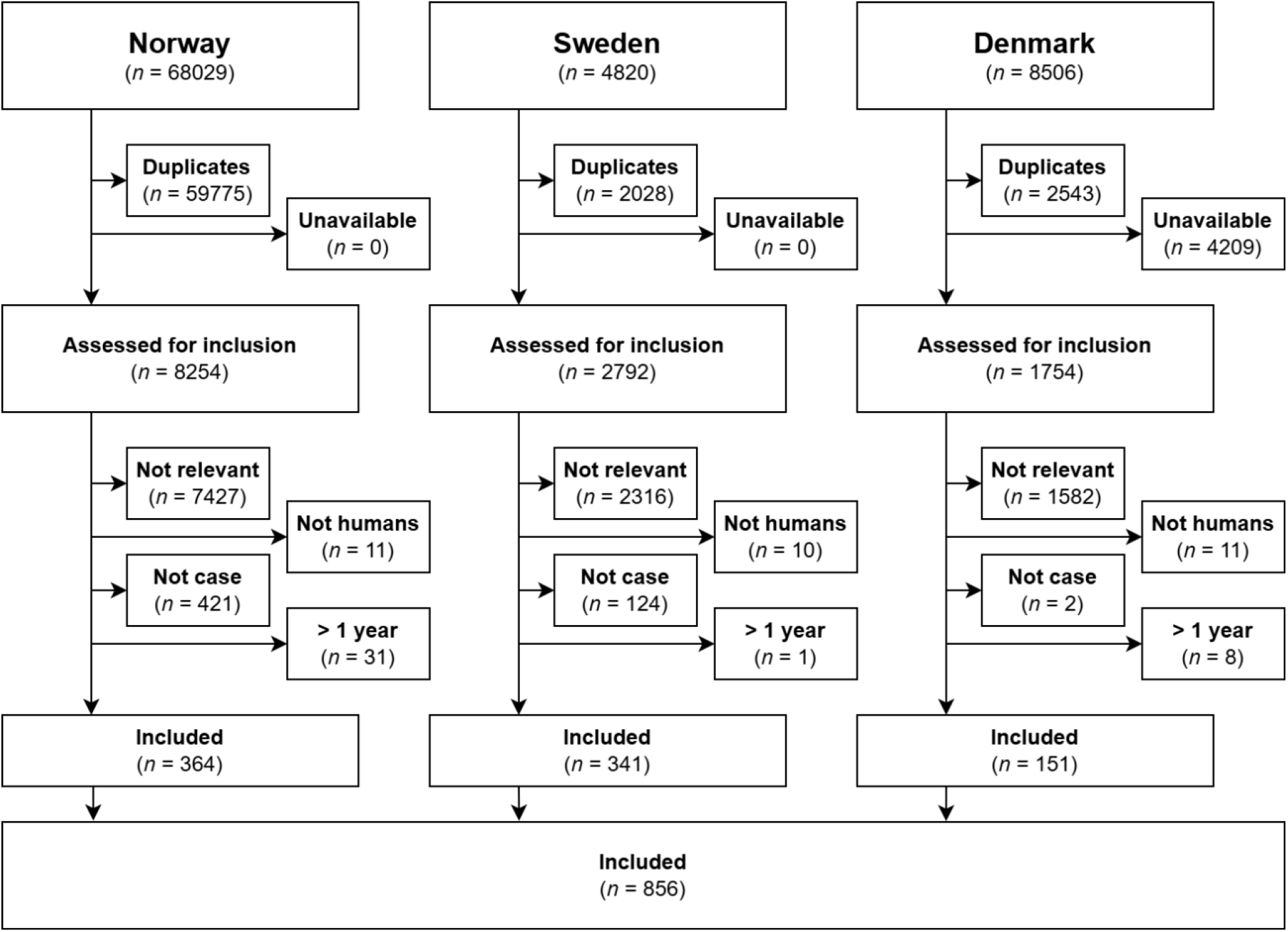
PRISMA flow chart for the selection of search items included in this systematic review from Norwegian, Swedish and Danish newspapers.

### Case overview

The earliest documented incidence occurred in 1845; a schoolboy in France froze his tongue to a metal bridge, causing loss of skin of tongue and lips as he forcefully detached himself (Case #1 in Table S2). The number of unique reported cases peaked during the 1950s before declining (Figure 2). Media coverage varied; most incidents (n=63, 56%) appeared in multiple newspapers and one case was mentioned in 76 different newspapers (Case #26 in Table S2). Eleven cases (10%) were reported across two countries, and one case (1%) appeared in newspapers from all three Scandinavian nations. The average (SD) time from the first to the last report of a single case was 17 (19) days.

**Figure 2.**
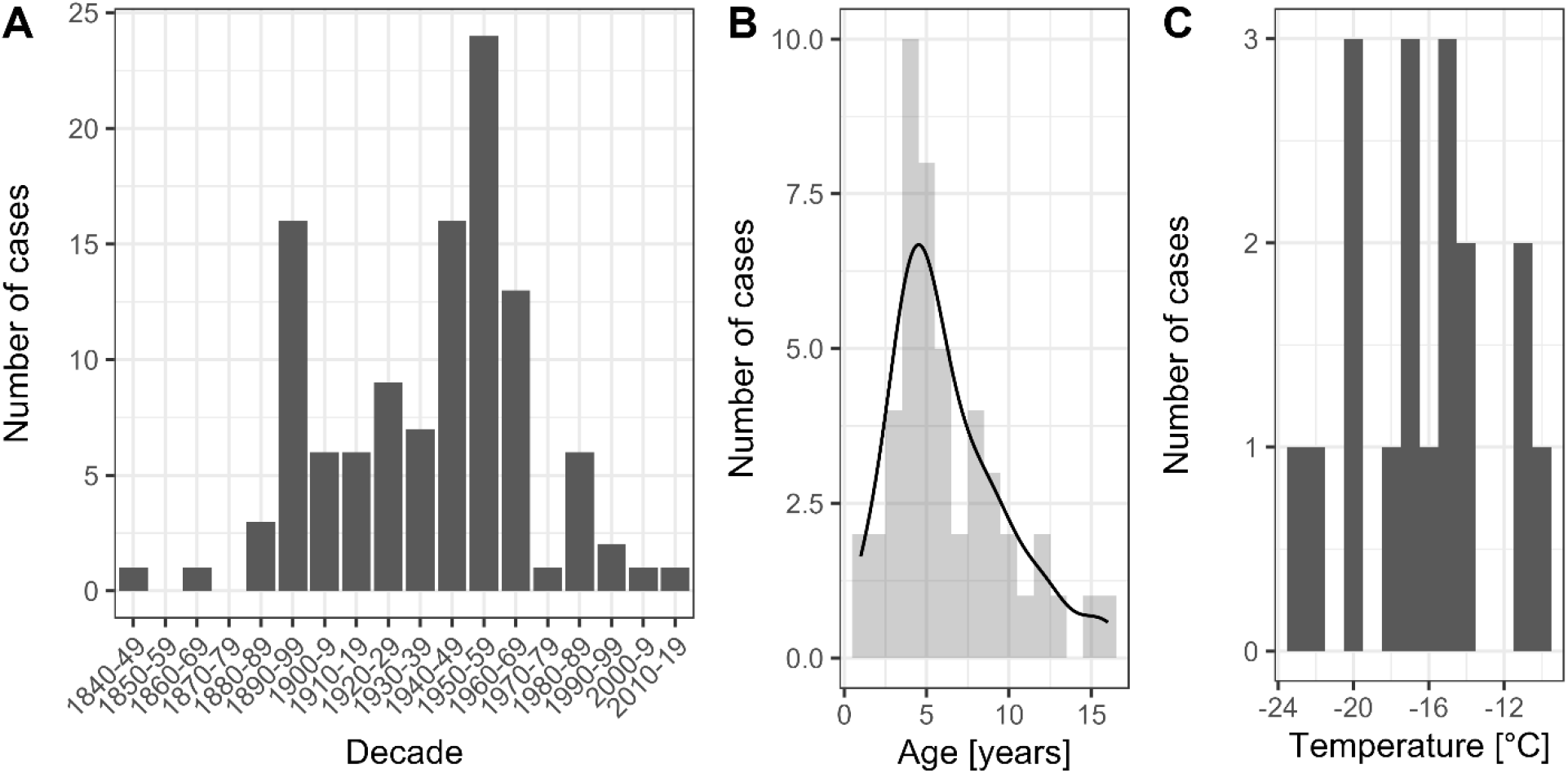
Number of cases, presented by (A) decade, (B) age of the involved person, and (C) ambient temperature at the incidence. Smoothed data distributions are shown as black lines in (B) and (C).

Almost all cases involved children (Table 1). Reported age ranged from 1 to 16 years (Fig. 2B) and ambient temperature ranged from -10 °C to -23 °C (Fig. 2C). Tongues adhered to various metal objects, most commonly railings (*n* = 45, 40%), fences (*n* = 15, 13%), lamp posts (*n* = 8, 7%), and, less commonly, objects such as a flagpole (*n* = 1, 1%) and a gold medal (*n* = 1, 1%). Most cases occurred outdoors in wintertime, except one case involving an indoor refrigeration system (Case #32 in Table S2) and a “ mass-casualty” involving children being served ice cream stored on dry ice causing laceration of the tongue (Case #107 in Table S2). There were multiple examples of copycat behavior; one case mentioned six children getting their tongue stuck after having read a story about it in the newspaper (Case #93 in Table S2).

**Table 1.** Case characteristics.

|  |  | **Not reported, N** |
| --- | --- | --- |
| Number of cases, N | 113 | — |
| Children (< 18 years of age), N (%) | 108 (96%) | 0 |
| Boys, N (%) | 71 (63%) | 7 |
| Age, years | 5.25 (1, 16) | 65 |
| Ambient temperature, °C | -16.5 (-23, -10) | 95 |
| Country, N (%) |  | 0 |
| Austria | 1 (1%) |  |
| Canada | 1 (1%) |  |
| Denmark | 18 (16%) |  |
| England | 4 (4%) |  |
| Finland | 3 (3%) |  |
| France | 2 (2%) |  |
| Germany | 1 (1%) |  |
| The Netherlands | 2 (2%) |  |
| Norway | 52 (46%) |  |
| Sweden | 27 (24%) |  |
| USA | 2 (2%) |  |
| Month of occurrence, N (%) |  | 47 |
| January | 26 (23%) |  |
| February | 13 (12%) |  |
| March | 7 (6%) |  |
| May | 1 (1%) |  |
| November | 6 (5%) |  |
| December | 13 (12%) |  |
| Involvement of, N (%) |  |  |
| Hospital | 9 (8%) | 104 |
| Doctor | 20 (18%) | 93 |
| Police | 8 (7%) | 105 |
| Fire service | 4 (4%) | 109 |

For categorical variables, percentage refers to total numbers. Continuous variables are reported as median (range).

Various remedies were employed to detach the frozen tongues, most often water (*n* = 34, 30%). Other remedies included glycerol, coffee, whiskey, a car cigarette lighter, a penknife, and heating with denatured alcohol. The longest reported time stuck was 90 minutes (Case #70 in Table S2).

The reported outcomes varied significantly in severity. At the mild end were outcomes such as pain, psychological distress and mild bleeding. However, seeking medical attention from a doctor or going to the hospital was not uncommon. The police or fire department was also frequently involved (Table 1). More serious consequences included hospitalization, potential amputation of tongue tissue, need for suturing, and possibly systemic infection. Another potentially serious incident involved a child getting stuck on the railway; luckily, he was discovered in time, and the approaching train was sent to another track (Case #40 in Table S2).

## Discussion

Despite being a recognized cultural phenomenon, tongue adhesion to cold metal (“ tundra tongue” ) has lacked systematic study. In this paper, we present the first overview of such cases based on more than 850 newspaper reports. Our findings document that potentially severe consequences could occur, although infrequently.

The demography of the study population was surprisingly diverse. Although only Scandinavian newspapers were examined, the incidents originated from 11 countries across two continents. The overrepresentation of boys aligns with previous research reporting higher injury rates among boys during outdoor play (23,24). The decrease in number of reports the last 50 years may represent an actual decrease in incidence, though this could be impacted by reporting bias and shifting media landscapes. Norway’s 1990s playground safety regulations are unlikely to be a major factor, as incidents typically involved non-playground objects like railings and lamp posts.

The dangers of the tundra tongue can manifest in at least three different ways. First, direct cold exposure can cause tissue injury. Ice crystal formation causes immediate tissue injury and is probable at tissue temperature -4.4 °C, and possible at tissue temperatures as high as -0.55 °C (25,26). Non-freezing cold injuries due to vasoconstriction and hypoperfusion occur only after several hours and are thus less relevant for the tundra tongue. White patches (e.g., Case #41 in Table S2) are generally considered a reversible sign of mild frostbite. In contrast, another case involved scarring in the face, described as potentially irreversible (Case #32 in Table S2). One case mentioned potential tissue amputation (Case #42 in Table S2), but the medical indication was not given. Another case serves as an extreme example of the damage of freezing (Case #11 in Table S2); a newspaper reported that a man fell asleep during a ski hike with his tongue exposed so that it was “ frozen stiff” when he woke up. Second, forceful detachment can cause mechanical trauma. Ulcers and bleeding were described in several cases. It is not known how deep these ulcers are and whether the depth of injury depends on factors such as metal temperature and contact time. Breach of the skin barrier increases the risk of infection; systemic symptoms of infection were reported in one case (Case #65 in Table S2). Third, getting stuck without the means to call for help or move can itself be dangerous. One child was stuck for 90 minutes, which might cause a significant risk for general hypothermia (Case #70 in Table S2). Getting stuck on the railway is another example of immediate and potentially life-threatening danger (Case #40 in Table S2). Interestingly, the latter reminds of an anecdote published only two years earlier (1927) in the same country (USA) describing a myth about a man getting beheaded after his tongue froze to a railway in Indiana (27, #1130).

Although most cases of the tundra tongue may not reach the newspapers, some evidently do. The selection of cases published in newspapers is likely to be biased and the population presented in this paper may thus differ significantly from the general tundra tongue population. Important limitations regarding the literature search include the accuracy of the library databases (especially older newspapers might have inaccurate transcriptions), wordings not covered by our searches, and incomplete library databases (of note, the Swedish collection is incomplete for the period 1907-2013). However, the observation that most identified incidents were reported across multiple newspapers somewhat lessens the impact of potentially missing individual news reports due to database or search imperfections.

## Conclusions

This scoping review of historical Scandinavian newspaper reports indicates that while most tundra tongue incidents appear mild and without long-term sequelae, severe injuries have occurred. Young children, particularly boys, represent the highest risk group.

Although ambient temperature data were sparse, no incidents were reported above -8 °C. Documented outcomes ranged widely, from minor distress and injury to hospital treatment and potentially permanent facial scarring. Therefore, parents, policymakers, and healthcare professionals should be aware of and not underestimate the potential harm associated with tundra tongue.

## Contributions

**AHJ**: Conceptualization, Methodology, Visualization, Project Administration, Writing – Original Draft Preparation, Formal Analysis, Investigation, Data Curation, Writing – Review & Editing. **SET**: Formal Analysis, Investigation, Data Curation, Validation, Writing – Review & Editing. **BCS**: Formal Analysis, Investigation, Data Curation, Validation, Writing – Review & Editing. **YH**: Formal Analysis, Validation, Writing – Review & Editing. **SHT**: Supervision, Writing – Review & Editing

## Acknowledgements

We would like to thank Henrik Døllner and Carina Ryssdal for their assistance in designing the Danish and Swedish search strategies.

## Data availability

The data that support the findings of this study are openly available in figshare at http://doi.org/10.6084/m9.figshare.28710698, reference10.6084/m9.figshare.28710698.

## Patient consent for publication

Not applicable.

## Ethics approval

Not applicable.

