## Supplementary material for "Demography and outcomes of frozen tongue: a scoping review of Scandinavian tundra tongue cases": Figure S1

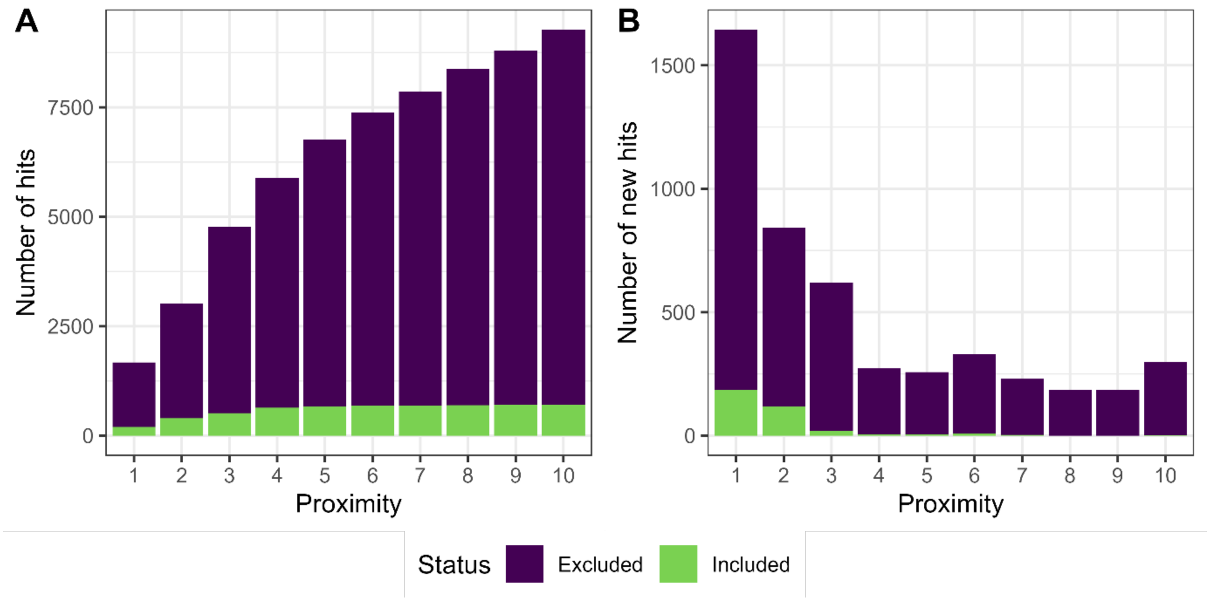


**Figure S1**. (A) The total number of hits and (B) the number of new hits as the maximum allowed distance (proximity) between two search terms was increased (Table S1). For example, using the two terms “tongue” and “stuck”, the phrase “the tongue got stuck” matches with proximity 1, whereas “the tongue immediately got stuck” matches with proximity 2.
