## Supplementary material for "Demography and outcomes of frozen tongue: a scoping review of Scandinavian tundra tongue cases": Figure S2

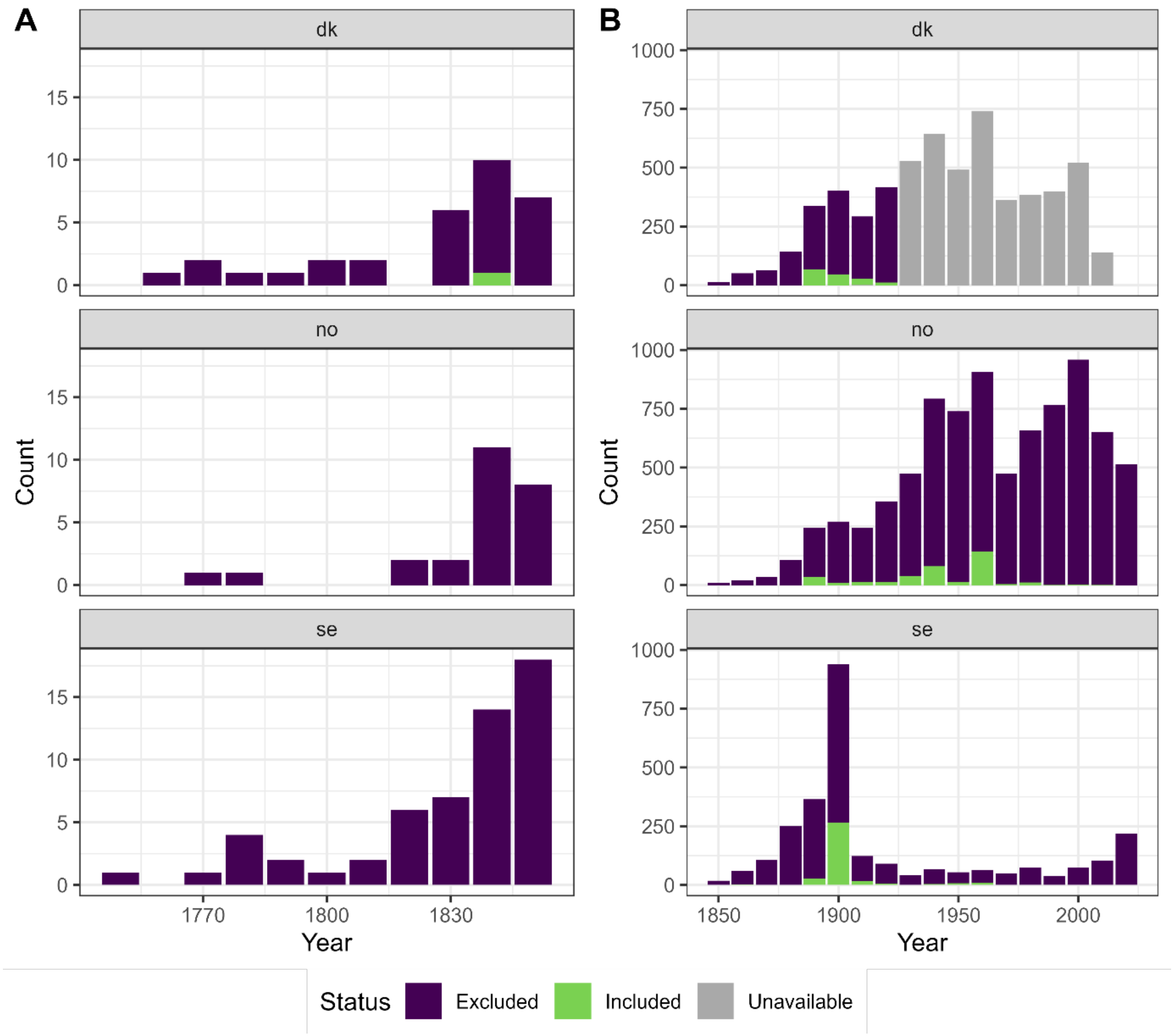


**Figure S2**. Number of search hits shown by year of publication (10-year bulks). The results have been divided into publication before (A) or after (B) January 1st, 1850, to better visualize the frequencies. Abbreviations: dk, Denmark; no, Norway; se, Sweden.
