## Supplementary material for "Demography and outcomes of frozen tongue: a scoping review of Scandinavian tundra tongue cases": Table S1

**Table S1**. Search terms and number of hits. The proportion of included items is based on the number of available items. The actual search terms in Norwegian, Swedish and Danish are given as notes.

| Search term | Hits (available) | Included |
|---|---|---|
| **Norwegian** | | |
| " stuck[1] tongue[2] "~3 | 46 (46, 100%) | 0 (0%) |
| " stuck[1] tongue[3] "~3 | 8 (8, 100%) | 0 (0%) |
| " stuck[1] tongen[4] "~3 | 10 (10, 100%) | 0 (0%) |
| " stuck[1] tongue[5] "~1 | 350 (350, 100%) | 9 (2.6%) |
| " stuck[1] tongue[5] "~2 | 502 (502, 100%) | 15 (3%) |
| " stuck[1] tongue[5] "~3 | 756 (756, 100%) | 32 (4.2%) |
| " stuck[1] tongue[5] "~4 | 965 (965, 100%) | 38 (3.9%) |
| " stuck[1] tongue[5] "~5 | 1162 (1162, 100%) | 42 (3.6%) |
| " stuck[1] tongue[5] "~6 | 1272 (1272, 100%) | 43 (3.4%) |
| " stuck[1] tongue[5] "~7 | 1392 (1392, 100%) | 44 (3.2%) |
| " stuck[1] tongue[5] "~8 | 1562 (1562, 100%) | 44 (2.8%) |
| " stuck[1] tongue[5] "~9 | 1670 (1670, 100%) | 44 (2.6%) |
| " stuck[1] tongue[5] "~10 | 1773 (1773, 100%) | 44 (2.5%) |
| " stuck[1] tongue[6] "~3 | 2021 (2021, 100%) | 15 (0.7%) |
| " stuck[1] tongue[7] "~1 | 652 (652, 100%) | 151 (23.2%) |
| " stuck[1] tongue[7] "~2 | 1279 (1279, 100%) | 268 (21%) |
| " stuck[1] tongue[7] "~3 | 1928 (1928, 100%) | 277 (14.4%) |
| " stuck[1] tongue[7] "~4 | 2080 (2080, 100%) | 278 (13.4%) |
| " stuck[1] tongue[7] "~5 | 2245 (2245, 100%) | 288 (12.8%) |
| " stuck[1] tongue[7] "~6 | 2402 (2402, 100%) | 298 (12.4%) |
| " stuck[1] tongue[7] "~7 | 2502 (2502, 100%) | 304 (12.2%) |
| " stuck[1] tongue[7] "~8 | 2630 (2630, 100%) | 304 (11.6%) |
| " stuck[1] tongue[7] "~9 | 2740 (2740, 100%) | 306 (11.2%) |
| " stuck[1] tongue[7] "~10 | 2888 (2888, 100%) | 307 (10.6%) |
| " tongue[2] stuck[1] "~3 | 48 (48, 100%) | 0 (0%) |
| " tongue[3] stuck[1] "~3 | 5 (5, 100%) | 0 (0%) |
| " tongue[4] stuck[1] "~3 | 4 (4, 100%) | 0 (0%) |
| " tongue[5] stuck[1] "~1 | 250 (250, 100%) | 23 (9.2%) |
| " tongue[5] stuck[1] "~2 | 561 (561, 100%) | 35 (6.2%) |
| " tongue[5] stuck[1] "~3 | 871 (871, 100%) | 40 (4.6%) |
| " tongue[5] stuck[1] "~4 | 989 (989, 100%) | 41 (4.1%) |
| " tongue[5] stuck[1] "~5 | 1152 (1152, 100%) | 44 (3.8%) |
| " tongue[5] stuck[1] "~6 | 1321 (1321, 100%) | 44 (3.3%) |
| " tongue[5] stuck[1] "~7 | 1463 (1463, 100%) | 44 (3%) |
| " tongue[5] stuck[1] "~8 | 1562 (1562, 100%) | 44 (2.8%) |
| " tongue[5] stuck[1] "~9 | 1653 (1653, 100%) | 44 (2.7%) |
| " tongue[5] stuck[1] "~10 | 1750 (1750, 100%) | 44 (2.5%) |
| " tongue[6] stuck[1] "~3 | 2105 (2105, 100%) | 20 (1%) |
| " tongue[7] stuck[1] "~1 | 417 (417, 100%) | 14 (3.4%) |

| | | |
|---|---|---|
| " tongue[7] stuck[1] "~2 | 674 (674, 100%) | 82 (12.2%) |
| " tongue[7] stuck[1] "~3 | 1210 (1210, 100%) | 168 (13.9%) |
| " tongue[7] stuck[1] "~4 | 1851 (1851, 100%) | 283 (15.3%) |
| " tongue[7] stuck[1] "~5 | 2211 (2211, 100%) | 292 (13.2%) |
| " tongue[7] stuck[1] "~6 | 2383 (2383, 100%) | 296 (12.4%) |
| " tongue[7] stuck[1] "~7 | 2506 (2506, 100%) | 299 (11.9%) |
| " tongue[7] stuck[1] "~8 | 2619 (2619, 100%) | 305 (11.6%) |
| " tongue[7] stuck[1] "~9 | 2726 (2726, 100%) | 307 (11.3%) |
| " tongue[7] stuck[1] "~10 | 2863 (2863, 100%) | 309 (10.8%) |
| **Swedish** | | |
| " stuck[1] tongue[8] "~3 | 1890 (1890, 100%) | 213 (11.3%) |
| " stuck[9] tongue[5] "~3 | 10 (10, 100%) | 7 (70%) |
| " stuck[9] tongue[8] "~3 | 97 (97, 100%) | 85 (87.6%) |
| " stuck[10] tongue[5] "~3 | 7 (7, 100%) | 0 (0%) |
| " stuck[10] tongue[8] "~3 | 66 (66, 100%) | 0 (0%) |
| " ice[11] licking[12] "~3 | 38 (38, 100%) | 0 (0%) |
| " cold[13] tongue[5] "~3 | 26 (26, 100%) | 0 (0%) |
| " cold[13] tongue[8] "~3 | 24 (24, 100%) | 1 (4.2%) |
| " iron[14] tongue[8] "~3 | 195 (195, 100%) | 45 (23.1%) |
| " licking[12] ice[11] "~3 | 40 (40, 100%) | 0 (0%) |
| " licking[12] bar[15] "~3 | 10 (10, 100%) | 0 (0%) |
| " bar[15] licking[12] "~3 | 1 (1, 100%) | 0 (0%) |
| " bar[15] tongue[5] "~3 | 29 (29, 100%) | 1 (3.4%) |
| " bar[15] tongue[8] "~3 | 6 (6, 100%) | 1 (16.7%) |
| " tongue[5] stuck[9] "~3 | 11 (11, 100%) | 7 (63.6%) |
| " tongue[5] stuck[10] "~3 | 5 (5, 100%) | 0 (0%) |
| " tongue[5] cold[13] "~3 | 33 (33, 100%) | 0 (0%) |
| " tongue[5] bar[15] "~3 | 29 (29, 100%) | 1 (3.4%) |
| " tongue[8] stuck[1] "~3 | 2010 (2010, 100%) | 270 (13.4%) |
| " tongue[8] stuck[9] "~3 | 100 (100, 100%) | 85 (85%) |
| " tongue[8] stuck[10] "~3 | 74 (74, 100%) | 0 (0%) |
| " tongue[8] cold[13] "~3 | 14 (14, 100%) | 1 (7.1%) |
| " tongue[8] iron[14] "~3 | 89 (89, 100%) | 32 (36%) |
| " tongue[8] bar[15] "~3 | 16 (16, 100%) | 2 (12.5%) |
| **Danish** | | |
| " stuck[1] tongue[6] "~3 | 2441 (479, 19.6%) | 7 (1.5%) |
| " stuck[1] tongue[7] "~3 | 1394 (581, 41.7%) | 86 (14.8%) |
| " tongue[6] stuck[1] "~3 | 2763 (611, 22.1%) | 34 (5.6%) |
| " tongue[7] stuck[1] "~3 | 1908 (931, 48.8%) | 133 (14.3%) |

*Notes:* [1]fast; [2]tonga; [3]tonge; [4]tongen; [5]tunga; [6]tunge; [7]tungen; [8]tungan; [9]fastfrusen; [10]fastklistrat; [11]is; [12]slicka; [13]iskall; [14]jarn; [15]stolpe
