## Supplementary material for "Demography and outcomes of frozen tongue: a scoping review of Scandinavian tundra tongue cases": Table S2

**Table S2.** Case overview and characteristics.

| # | Date, Location | Age, Sex | °C | Object | Remedy | Outcome | Involvement | References |
|---|---|---|---|---|---|---|---|---|
| 1 | 1845<br>France | Boy | | Iron railing | | Skin segments were avulsed from the tongue and lips | | 1 |
| 2 | 1861<br>France | Man | | Pump rod | Boiling water | A skin segment was avulsed from the tongue | | 2-3 |
| 3 | January 1888<br>Finland | Girl | | Brass railing | | | | 4-6 |
| 4 | March 1888<br>Norway | Girl | -17 | Iron railing | Warm water | Bleeding from the tongue | | 7-41 |
| 5 | March 1888<br>Sweden | Girl | | Iron railing | Warm water | Bleeding from the tongue | | 42-65 |
| 6 | January 1891<br>Denmark | Boy | | Door handle | Hit by his mother | A piece of tissue was avulsed from the tongue, extensive bleeding | | 66-68 |
| 7 | 1892<br>Denmark | Man | -11 | Iron railing | Warm water | Unable to speak for a short time | | 69-102 |
| 8 | 1893<br>Denmark | Boy | | Lamp post | | A skin segment was avulsed from the tongue | | 103-107 |
| 9 | 1893<br>Denmark | Boy | | Iron railing | | Impaired speech for a short time | | 108-131 |
| 10 | 1894<br>Sweden | Girl | | Piece of iron | | Extensive injury of the tongue | | 132-156 |
| 11 | February 1895<br>Sweden | Man | | * | | The tongue was frozen, underwent surgery to regain speech | H D | 150 |
| 12 | February 1895<br>Denmark | 10-15y<br>Girl | | Iron fence | | A skin segment and a piece of tissue were avulsed from the tongue. Treated by doctor | H D | 157-168 |
| 13 | 1895<br>Finland | Boy | | Exterior door lock | | A skin segment was avulsed from the tongue | | 169 |

| # | Date, Location | Age, Sex | °C | Object | Remedy | Outcome | Involvement | References |
|---|---|---|---|---|---|---|---|---|
| 14 | 1895<br>England | Boy | | Iron fence | | Skin segments were avulsed from the tongue | | 170-177 |
| 15 | February 1895<br>Norway | Girl | | Iron railing | | A skin segment was avulsed from the tongue | | 178-181 |
| 16 | 1895<br>England | Girl | | Iron fence | | | | 170-177 |
| 17 | February 1895<br>Denmark | Boy | | Iron railing | His sister forcefully pulled him away | A skin segment was avulsed from the tongue | | 182-186 |
| 18 | February 1897<br>Norway | 5-10y<br>Boy | | Water source | Warm water | Pain | | 187-191 |
| 19 | January 1898<br>Sweden | 10-15y<br>Girl | -18 | Gate railing | By herself | Bleeding from lips and tongue | | 192-263 |
| 20 | November 1898<br>Sweden | Girl | | Iron railing | | The tongue was badly injured | | 264-271 |
| 21 | 1899<br>Sweden | Boy | | Iron bar | Had to be forcibly detached from the iron | Bleeding from the tongue | | 272 |
| 22 | 1901<br>Denmark | Boy | | Iron railing | Lukewarm water | Discomfort | | 273-283 |
| 23 | 1902<br>Sweden | 5-10y<br>Girl | | Iron railing | | A skin segment was avulsed from the tongue | | 284-295 |
| 24 | November 1903<br>Denmark | Girl | | Lamp post | | A skin segment was avulsed from the tongue, severe pain | | 296-329 |
| 25 | 1903<br>Sweden | 15-20y<br>Boy | | Piece of iron | Metal heated by another hot piece of metal | Bleeding from the tongue | | 330-392 |
| 26 | December 1905<br>Sweden | Boy | | Railway fence | Metal heated by hands | Multiple skin segments were avulsed from the tongue and the mouth, extensive bleeding | H D | 393-466 |
| 27 | January 1907<br>Norway | Girl | | Iron railing | | Bleeding from the tongue | D | 467-474 |

| # | Date, Location | Age, Sex | °C | Object | Remedy | Outcome | Involvement | References |
|---|---|---|---|---|---|---|---|---|
| 28 | 1910<br>Norway | Girl | | Railing | A policeman heated the metal | | P | 475-479 |
| 29 | January 1912<br>Sweden | < 5y<br>Boy | -15 | Lamp post | Metal heated by hands | Almost no injury | | 480-484 |
| 30 | February 1912<br>Denmark | < 5y<br>Boy | | Iron railing | Warm water | No sequelae | | 485-491 |
| 31 | February 1912<br>Denmark | Boy | | Pump rod | Warm water | Multiple skin segments were avulsed from the tongue | | 489, 492-510 |
| 32 | 1914<br>USA | 10-15y<br>Girl | | Ammonia piping in refrigeration system | | Complete avulsion of a tissue segment. Significant facial frostbite lesion, with potential for persistent scarring | | 511 |
| 33 | 1915<br>Sweden | 5-10y<br>Boy | | Iron fence | Assistance from policeman | A skin segment was avulsed from the tongue | P | 512 |
| 34 | 1920<br>Norway | Girl | | Brass railing | Metal heated by the respiration of two adults | Bleeding from the tongue | | 513-522 |
| 35 | 1920<br>Sweden | < 5y<br>Boy | | Iron guardrail | Assistance from policeman | | P | 523-526 |
| 36 | 1923<br>Denmark | **<br>Boy | | Iron bar | Warm water | Extensive bleeding from the tongue | | 527-528 |
| 37 | December 1923<br>Denmark | Boy | | Lamp post | Warm water | Bleeding from the tongue, pain | | 529-539 |
| 38 | 1926<br>Norway | | | Water pump | | A skin segment was avulsed from the tongue | | 540 |
| 39 | 1927<br>Denmark | Boy | | Iron railing | | Extensive bleeding from the tongue | | 541-554 |
| 40 | December 1929<br>USA | 15-20y<br>Boy | | Steel rail on railway premises | Steam was directed onto the railway rail | He heard an approaching locomotive and succeeded in attracting the attention of | | 555-556 |

| # | Date, Location | Age, Sex | °C | Object | Remedy | Outcome | Involvement | References |
|---|---|---|---|---|---|---|---|---|
| | | | | | | railway personnel; locomotive switched to adjacent track | | |
| 41 | January 1929 Norway | 5-10y Boy | -20 | Railing | The metal was heated by the boy's own respiration | White patch on the tongue | | 557 |
| 42 | 1929 Austria | Boy | | Iron grating | Firemen came and heated the metal | The child hospitalized, a portion of the tongue may require amputation | H D F | 558-571 |
| 43 | November 1930 Norway | | | Iron railing | Warm water | | | 572 |
| 44 | March 1931 Norway | Boy | | Railing | The metal was heated | | | 573-578 |
| 45 | January 1936 Norway | Boy | -23 | Iron railing | Warm water | No sequelae | | 579-592 |
| 46 | 1936 England | Boy | | Iron railing | Glycerin | | | 593-598 |
| 47 | December 1937 Sweden | 5-10y Boy | | Iron railing | Warm water | Bleeding from the tongue | | 599 |
| 48 | December 1937 Norway | < 5y Boy | -10 to -12 | Iron rod | | A skin segment was avulsed from the tongue | | 600 |
| 49 | 1938 Norway | 5-10y Boy | | Axe hammer | Warm water | | | 601-603 |
| 50 | January 1940 Denmark | Girl | | Iron railing | Help from older boy | A piece of tissue was avulsed from the tongue, extensive bleeding | | 604-605 |
| 51 | 1941 Norway | | | Iron object | | No sequelae | | 606 |
| 52 | January 1941 Norway | 5-10y Boy | | Iron gate | Metal heated by hands | No sequelae | | 606-628 |

| # | Date, Location | Age, Sex | °C | Object | Remedy | Outcome | Involvement | References |
|---|---|---|---|---|---|---|---|---|
| 53 | 1941<br>Norway | Boy | | Iron fence | A man warmed the iron with his hands | | | 629 |
| 54 | 1941<br>Norway | Girl | | Fence post | Detached herself by force | A skin segment was avulsed from the tongue | | 630-633 |
| 55 | January 1942<br>Sweden | 10-15y<br>Boy | | Iron fence | Cloth with warm water applied by ambulance personel | Lacerations of the tongue and lower lip, no permanent sequelae | D | 634-635 |
| 56 | 1942<br>Denmark | < 5y<br>Girl | | Iron railing | Adults helped her detach with a jerk motion | Lacerations of the tongue, extensive bleeding, brought to hospital by ambulance | H D | 636-644 |
| 57 | March 1942<br>Denmark | 10-15y<br>Girl | | Lamp post | Warm water | Not severe injury, but brought to hospital | H D | 645-658 |
| 58 | 1945<br>Norway | Boy | | Iron railing | Warm water | Bleeding from the tongue, paresthesia | | 659-662 |
| 59 | 1946<br>Norway | Boy | | Iron railing | Warm water | No sequelae | | 663 |
| 60 | 1946<br>Norway | Boy | | Woven wire fence | | A skin segment was avulsed from the tongue | | 664 |
| 61 | December 1946<br>Sweden | Boy | | Iron fence | A nurse warmed the iron with her hands | Skin segments were avulsed from the tongue, no permanent sequelae | | 665 |
| 62 | February 1947<br>Sweden | Girl | -17 | Iron railing | | A piece of tissue was avulsed from the tongue, extensive bleeding | | 666 |
| 63 | March 1947<br>Sweden | < 5y<br>Boy | | Piece of iron ("järnstag") | Warm water | Skin segments were avulsed from the tongue, pain and discomfort | | 667 |
| 64 | January 1948<br>Norway | < 5y<br>Girl | | Railing | | Extensive bleeding | | 668 |
| 65 | January 1949<br>Norway | Girl | | Metal object | | The entire skin of the tongue was avulsed. | D | 669 |

| # | Date, Location | Age, Sex | °C | Object | Remedy | Outcome | Involvement | References |
|---|---|---|---|---|---|---|---|---|
| | | | | | | Repeated doctor treatments. For a period, her condition deteriorated significantly, accompanied by a high-grade fever. In recovery | | |
| 66 | January 1950<br>Sweden | < 5y<br>Boy | | Iron railing | Detached himself by force | Bleeding from the tongue | | 670 |
| 67 | 1951<br>Norway | 5-10y<br>Boy | | Iron bar | Warm water | Bleeding from the tongue and lips | | 671-672 |
| 68 | January 1952<br>Sweden | < 5y<br>Boy | -14 | Lamp post | Cold water applied by police | Exhaution and distress | P | 673-675 |
| 69 | January 1953<br>Norway | Boy | | Metal handle on a railway carriage | [7 min.] | | | 676 |
| 70 | November 1954<br>Norway | 5-10y<br>Boy | -14 to -15 | Iron bar | [90 min.] | | | 677-682 |
| 71 | December 1955<br>Sweden | Boy | | Iron fittings | Warm water | A skin segment was avulsed from the tongue | | 683 |
| 72 | December 1955<br>Norway | Girl | | Iron railing | Car cigarette lighter | No sequelae, pain | | 684-685 |
| 73 | January 1955<br>Norway | Boy | | Iron bar | Detached himself by force | A skin segment was avulsed from the tongue | | 686-697 |
| 74 | 1955<br>Germany | 5-10y<br>Boy | | Iron railing | A doctor used almost an hour to detach him | | D | 698 |
| 75 | December 1955<br>Norway | 5-10y<br>Boy | | Iron railing | Assistance from policeman | Skin segmenst were avulsed from the tongue, bleeding. No serious injury | P | 699-770 |
| 76 | 1955<br>Norway | < 5y<br>Girl | | Metal on fence | A teacher warmed the iron with his/her hands | | | 771 |
| 77 | February 1956<br>The Netherlands | Girl | | Iron railing | Hot coffee | | | 772 |

| # | Date, Location | Age, Sex | °C | Object | Remedy | Outcome | Involvement | References |
|---|---|---|---|---|---|---|---|---|
| 78 | February 1956<br>Norway | Boy | | Iron fence | | No sequelae | | 773 |
| 79 | January 1956<br>Sweden | 5-10y<br>Boy | | Marble slab | Hair dryer and warm water | Trouble eating for a few days | D | 774-775 |
| 80 | January 1956<br>Norway | 5-10y<br>Boy | | | | Mild bleeding from the tongue | | 776 |
| 81 | 1956<br>Denmark | 5-10y<br>Boy | | Iron railing | Detached himself by force | A skin segment was avulsed from the tongue | D | 777 |
| 82 | January 1958<br>Norway | | | Railing | | | | 778 |
| 83 | 1957<br>Denmark | 10-15y<br>Boy | | Flagpole | Detached himself by force [10 min.] | Skin segmenst were avulsed from the tongue | D | 779-781 |
| 84 | February 1958<br>Norway | 5-10y<br>Boy | | Snowplow | | Injury of parts of the tongue, bleeding | | 782-784 |
| 85 | January 1958<br>Norway | < 5y<br>Girl | | Iron railing | Detached herself by force | Laceration of the tongue, sutured by doctor | D | 785 |
| 86 | December 1958<br>Sweden | Boy | | Iron railing | Warm water | | | 786 |
| 87 | December 1958<br>Canada | 10-15y<br>Girl | | Iron railing | Whiskey applied by a policeman | | P | 787-816 |
| 88 | February 1959<br>Norway | Girl | | Iron bar | Warm water | Laceration of the tongue | | 817 |
| 89 | January 1959<br>Norway | < 5y<br>Girl | | Statue | Warm water | Laceration of the tongue. The chin was also exposed to cold. Immediate medical attention was required. The injury was not severe | D | 818-823 |
| 90 | 1960<br>Norway | Girl | | Woven wire fence | Warm water | | | 824 |

| # | Date, Location | Age, Sex | °C | Object | Remedy | Outcome | Involvement | References |
|---|---|---|---|---|---|---|---|---|
| 91 | January 1960 Sweden | 5-10y Boy | -12 to -15 | Lamp post | A doctor dissected the tongue free with a penknife | Mild bleeding from the tongue | D | 825 |
| 92 | 1960 Norway | 5-10y Boy | | Iron bar | Assitance from adults | No sequelae | | 826 |
| 93 | 1961 The Netherlands | Boy | | Steel plate | Warm water. Assistance from firemen | | F | 827 |
| 94 | March 1962 Norway | Boy | | Iron railing | Cloth with warm water | A skin segment was avulsed from the tongue, bleeding | | 828 |
| 95 | 1963 Norway | | | Garden fence | Needed assistance | A skin segment was avulsed from the tongue, pain | | 829 |
| 96 | November 1965 Norway | Boy | -19.5 | Railing | | | | 830-831 |
| 97 | November 1965 Sweden | < 5y Boy | -15 | Iron bar | | A skin segment was avulsed from the tongue, extensive bleeding. Brought to hospital, no permanent sequelae | H D | 832 |
| 98 | December 1965 Sweden | 5-10y Boy | | Iron railing | Denatured alcohol was ignited beneath the iron bar. Ambulance, police, and fire department were summoned | | D P F | 833 |
| 99 | 1966 Norway | < 5y Boy | | Railing | His father forcefully pulled him away | | | 834 |
| 100 | 1966 Sweden | Boy | | Iron railing | Cloth with warm water. Assistance from policeman | Almost no injury, served cake by the police | P | 835 |
| 101 | February 1966 Norway | Boy | | Bronze statue | | | | 836 |

| # | Date, Location | Age, Sex | °C | Object | Remedy | Outcome | Involvement | References |
|---|---|---|---|---|---|---|---|---|
| 102 | 1968<br>Norway | Girl | | Fence | Lukewarm water | | | 837-838 |
| 103 | January 1977<br>Norway | 5-10y<br>Boy | -22 | Statue | Warm water | A skin segment was avulsed from the tongue | | 839-841 |
| 104 | January 1981<br>Norway | 5-10y<br>Girl | -17 | Post | Spit and jerk | A skin segment was avulsed from the tongue, extensive bleeding | H D | 842-844 |
| 105 | 1982<br>Norway | | | Iron pile | Lukewarm water | White patch on the chin. Frostbite injury to the tongue | | 845-846 |
| 106 | January 1984<br>Norway | 5-10y<br>Boy | | Railing | Warm water | Extensive bleeding from the tongue | | 847 |
| 107 | May 1984<br>Norway | ** | | Ice cream | Assistance from parents | Laceration of the tongue | | 848 |
| 108 | March 1984<br>Norway | < 5y<br>Girl | | Rear fender of car | | Bleeding from the tongue | | 849 |
| 109 | December 1986<br>Norway | Man | | Photo camera | | | | 850 |
| 110 | 1999<br>England | 5-10y<br>Boy | | Freezer | Hair dryer, assistance from fire department | | F | 851 |
| 111 | January 1999<br>Sweden | < 5y<br>Girl | -20 | Lamp post | His mother forcefully pulled her away | Bleeding from the tongue, with potential for taste impairment | H D | 852 |
| 112 | NR 2001<br>Finland | Man | -15 to -17 | Gold medal | | | | 853-854 |
| 113 | NR 2010<br>Norway | < 5y<br>Girl | -8 to -12 | Metal bar | Jerk | Bleeding from the tongue, pain | | 855-856 |

*Notes:* * The man fell asleep with his tongue exposed so that it froze, but it did not adhere to anything. ** The case involves several children.

*Abbreviations:* D, Doctor; F, Fire Service; H, Hospital; P, Police.

2. Nya Dagligt Allehanda [Internet]. 1861 Feb 04:3. Available from: https://tidningar.kb.se/6qjx674j55r1mz8/part/1/page/3

3. Norrländska korrespondenten [Internet]. 1861 Mar 09:3. Available from: https://tidningar.kb.se/h1t6mftt55w7x6g/part/1/page/3

4. Nya Tiden (Kristianstad : 1888) [Internet]. 1888 Jan 28:3. Available from: https://tidningar.kb.se/n13nw5czlgjtltcd/part/1/page/3

5. Stockholms nyheter [Internet]. 1888 Jan 26:3. Available from: https://tidningar.kb.se/t9jfn7jrr08vvbc2/part/1/page/3

6. Nya Dagligt Allehanda [Internet]. 1888 Jan 24:3. Available from: https://tidningar.kb.se/tc5kkxd53b4gl8m/part/1/page/3

7. Bergens Aftenblad [Internet]. 1888 Mar 20:2. Available from: https://www.nb.no/items/0194102c1cfb54a6fc74a22ffe582f0e

8. Ugeblad for Frederikstad og omegn [Internet]. 1888 Mar 26:1. Available from: https://www.nb.no/items/03335472091205980e659cc9968034aa

9. Thisted Amts Tidende (1882-1970) [Internet]. 1888 Mar 23:2. Available from: http://hdl.handle.net/109.3.1/uuid:12b223ce-6acc-496a-8ce2-b32557c1c7f5

10. Søndre Bergenhus Folkeblad [Internet]. 1888 Mar 28:2. Available from: https://www.nb.no/items/159863386f14970828351dcfb2062f16

11. Vestfold [Internet]. 1888 Mar 20:2. Available from: https://www.nb.no/items/192edf83479ea258afe69e6b17c97e64

12. Bratsberg Amtstidende [Internet]. 1888 Mar 23:2. Available from: https://www.nb.no/items/2ccc51e8a9d96a7a1613f63eedda2616

13. Fredericia Dagblad (1881-1890) [Internet]. 1888 Mar 22:2. Available from: http://hdl.handle.net/109.3.1/uuid:367a43ae-555b-453a-9a55-24069ade18d8

14. Kongsberg Blad [Internet]. 1888 Mar 24:2. Available from: https://www.nb.no/items/37b14a0a344adcc93b37819c03113bbe

15. Aalborg Stiftstidende (1885-1901) [Internet]. 1888 Mar 23:3. Available from: http://hdl.handle.net/109.3.1/uuid:3d718556-283f-4896-b3ae-48c2d533ce99

16. Grenmar [Internet]. 1888 Mar 15:2. Available from: https://www.nb.no/items/41a2b74f5b36413d4772ea9e1be701f0

17. Samhold (Gjøvik: 1885-1920) [Internet]. 1888 Mar 20:2. Available from: https://www.nb.no/items/47468f1ab42a8154c6c2f347d198342b

18. Næstved Tidende. Sydsjællands Folkeblad (1866-1918) [Internet]. 1888 Mar 24:3. Available from: http://hdl.handle.net/109.3.1/uuid:58bb254c-0b8c-466c-8795-105655f12b2c

19. Moss Tilskuer [Internet]. 1888 Mar 24:2. Available from: https://www.nb.no/items/5ce46e4fc91687ad8da4184515919b43

20. Folketidende [Internet]. 1888 Mar 17:2. Available from: https://www.nb.no/items/607069f5fb6d74650a6095997cbdc848

21. Thisted Amtsavis (1859-1960) [Internet]. 1888 Mar 23:2. Available from: http://hdl.handle.net/109.3.1/uuid:65e0ac84-baa8-454f-ac31-54309a583077

22. Jarlsberg og Larviks Amtstidende [Internet]. 1888 Mar 16:2. Available from: https://www.nb.no/items/757a72b3b1208b07e8d8c953a5610fc8

23. Lister og Mandals Amtstidende og Adresseavis [Internet]. 1888 Mar 24:2. Available from: https://www.nb.no/items/79f4a1ce88798ef49b395f17bca9d9ae

24. Horsens Folkeblad (1866-) [Internet]. 1888 Mar 23:2. Available from: http://hdl.handle.net/109.3.1/uuid:8211761b-2cf0-40b5-a2b0-9ff0cee4206f

25. Flekkefjords Budstikke [Internet]. 1888 Mar 21:2. Available from: https://www.nb.no/items/82e500a14202f124f022b0634c78297f

26. Tunsbergeren [Internet]. 1888 Mar 20:2. Available from: https://www.nb.no/items/87b385ee88ac27698365edb147b9c672

27. Grimstad-posten [Internet]. 1888 Mar 24:2. Available from: https://www.nb.no/items/930d5ae6f416f84fdee32a082f6acb7b

28. Drammens Tidende (Drammen: 1845-1960) [Internet]. 1888 Mar 16:2. Available from: https://www.nb.no/items/97486ab22a70427c4ec6c32512508613

29. Fremskridt [Internet]. 1888 Mar 17:2. Available from: https://www.nb.no/items/9d340bbd21b3956e63efb9420c15865e

30. Morgengryet [Internet]. 1888 Mar 17:2. Available from: https://www.nb.no/items/a667580395039597c0563a02ef33a737

31. Vestmar [Internet]. 1888 Mar 20:4. Available from: https://www.nb.no/items/b6b2e4b44686f861b8a4adacded6ee74

32. Morgenbladet [Internet]. 1888 Mar 15:2. Available from: https://www.nb.no/items/bd5e48391086c4f9360cad3e13d52236

33. Fjordenes Blad [Internet]. 1888 May 30:4. Available from: https://www.nb.no/items/bfce516019403ce403d2562ce3b68223

34. Kristianssands Stiftsavis og Adresse-Contors Efterretninger [Internet]. 1888 Mar 22:2. Available from: https://www.nb.no/items/c10aca6e52f80f635754783a46a14199

35. Kongsbergeren [Internet]. 1888 Mar 21:. Available from: https://www.nb.no/items/c157c800841237b06a496e6cb7aaa74f

36. Nordre Trondhjems Amtstidende [Internet]. 1888 Mar 20:2. Available from: https://www.nb.no/items/c16f4e2ae85cfc9c438e8120fc05544d

37. Stavangeren [Internet]. 1888 Mar 20:2. Available from: https://www.nb.no/items/e00b684f0129d76d10db436db7a7f970

38. Herning Folkeblad - Vestjylland (1883-) [Internet]. 1888 Mar 27:2. Available from: http://hdl.handle.net/109.3.1/uuid:e7095c83-ca0d-4a13-a99d-b114448c3b33

39. Agder [Internet]. 1888 Mar 20:2. Available from: https://www.nb.no/items/ef35759855654240210b3cf4d2d717af

40. Adressetidende for Brevig, Stathelle, Langesund, Bamble og Eidanger [Internet]. 1888 Mar 20:2. Available from: https://www.nb.no/items/f3f7dbde377ea812490ad7133e590671

41. Agderposten [Internet]. 1888 Mar 17:2. Available from: https://www.nb.no/items/f6c56d812c1230a94c40bb68640900db

42. Enköpings tidning (1885) [Internet]. 1888 Mar 22:3. Available from: https://tidningar.kb.se/1gq73fbkzl3r63wl/part/1/page/3

43. Falköpings tidning [Internet]. 1888 Mar 17:3. Available from: https://tidningar.kb.se/3mfshq0f3wl41jp/part/1/page/3

44. Nya Wermlandstidningen [Internet]. 1888 Mar 15:3. Available from: https://tidningar.kb.se/3mft6qhf0xz5lcr/part/1/page/3

45. Wermländingen [Internet]. 1888 Mar 21:3. Available from: https://tidningar.kb.se/4jrkcbzl2lpg84cz/part/1/page/3

46. Västgöta korrespondenten Skövde tidning [Internet]. 1888 Mar 16:3. Available from: https://tidningar.kb.se/5jd82ddx33s9rzhj/part/1/page/3

47. Karlshamn [Internet]. 1888 Mar 21:2. Available from: https://tidningar.kb.se/5l1nzf8h37fsjbg7/part/1/page/2

48. Malmö handels- och sjöfartstidning [Internet]. 1888 Mar 17:3. Available from: https://tidningar.kb.se/8lq73bkx6vbsrl1l/part/1/page/3

49. Skånska aftonbladet [Internet]. 1888 Mar 16:3. Available from: https://tidningar.kb.se/8qhjhxcg6m3d7hlc/part/1/page/3

50. Gefleposten (1864) [Internet]. 1888 Mar 20:3. Available from: https://tidningar.kb.se/bsjgj55q8xs10s6d/part/1/page/3

51. Ystads allehanda [Internet]. 1888 Mar 20:3. Available from: https://tidningar.kb.se/ct2qmgsd9b2xbgc3/part/1/page/3

52. Tidning för Falu län och stad [Internet]. 1888 Mar 17:3. Available from: https://tidningar.kb.se/gthr442qdwmkgr3c/part/1/page/3

53. Stockholms läns tidning (1886) [Internet]. 1888 Mar 16:4. Available from: https://tidningar.kb.se/kzdbdbcmhn6wbjvr/part/1/page/4

54. Ljungbyposten (1878) [Internet]. 1888 Mar 16:2. Available from: https://tidningar.kb.se/lzf6svvvjr5hzh9r/part/1/page/2

55. Folkets tidning [Internet]. 1888 Mar 16:2. Available from: https://tidningar.kb.se/m0zhqw52kpzhrtvq/part/1/page/2

56. Jämtlandsposten [Internet]. 1888 Mar 21:3. Available from: https://tidningar.kb.se/m5zd912z4b1c3cs/part/1/page/3

57. Helsingborgstidningen Skånes allehanda (1881) [Internet]. 1888 Mar 20:3. Available from: https://tidningar.kb.se/n1bnpbg0lw9vwv93/part/1/page/3

58. Smålands allehanda [Internet]. 1888 Mar 14:3. Available from: https://tidningar.kb.se/n399hgvql4j3d892/part/1/page/3

59. Blekinge läns tidning [Internet]. 1888 Mar 17:2. Available from: https://tidningar.kb.se/p1ckgw0pm8x6176x/part/1/page/2

60. Westmanlands allehanda [Internet]. 1888 Mar 16:3. Available from: https://tidningar.kb.se/p3c1g0phmtn60s44/part/1/page/3

61. Östgöten (Linköping : 1874) [Internet]. 1888 Mar 14:4. Available from: https://tidningar.kb.se/q26fgj59npfkrwjh/part/1/page/4

62. Södra Dalarnes tidning [Internet]. 1888 Mar 20:3. Available from: https://tidningar.kb.se/r4mxc4m3pxdtr13m/part/1/page/3

63. Smålänningen (Växjö : 1885) [Internet]. 1888 Mar 20:3. Available from: https://tidningar.kb.se/t9xtvvg8r89qp9cx/part/1/page/3

64. Lindesposten (Göteborg : 1879) [Internet]. 1888 Mar 21:3. Available from: https://tidningar.kb.se/w8m1z8mdts98njs3/part/1/page/3

65. Gefleposten veckoupplagan [Internet]. 1888 Mar 22:4. Available from: https://tidningar.kb.se/wc3vmn9xtl88lqfb/part/1/page/4

66. Thisted Amtsavis (1859-1960) [Internet]. 1891 Jan 23:2. Available from: http://hdl.handle.net/109.3.1/uuid:3e961424-d307-4c7e-92af-43896d4ce6d4

67. Randers Amtsavis og Adressecontoirs Efterretninger (1857-1947) [Internet]. 1891 Jan 22:3. Available from: http://hdl.handle.net/109.3.1/uuid:bceae1e9-f71e-4f0e-9311-e68f2c386c35

68. Aarhuus Stifts-Tidende (1871-1989) [Internet]. 1891 Jan 24:1. Available from: http://hdl.handle.net/109.3.1/uuid:f9630f80-8ff4-4720-8807-86a097ad4c9b

69. Kongsberg Blad  [Internet]. 1892 Jan 20:2. Available from: https://www.nb.no/items/05d37fdfbf150a795715643a9be4dad1

70. Sjællands-Posten (Ringsted) (1853-1906) [Internet]. 1892 Jan 12:2. Available from: http://hdl.handle.net/109.3.1/uuid:121162c3-dd9b-4435-9514-fdd47319d94c

71. Thisted Amtsavis (1859-1960) [Internet]. 1892 Jan 11:2. Available from: http://hdl.handle.net/109.3.1/uuid:19b739d1-199b-4c27-8c7a-f7fab8108070

72. Randers Dagblad og Folketidende (1874-1970) [Internet]. 1892 Jan 12:2. Available from: http://hdl.handle.net/109.3.1/uuid:2a3212b7-c4ef-4198-8342-a51032d9d1e1

73. Vejle Amts Folkeblad (1865-) [Internet]. 1892 Jan 13:2. Available from: https://www2.statsbiblioteket.dk/mediestream/avis/record/doms_aviser_page:uuid:2cd3e1bb-49db-466f-95cf-bbe6ff2d1d84/query/%22tungen%20fast%22~3

74. Holmestrandsposten  [Internet]. 1892 Jan 16:2. Available from: https://www.nb.no/items/310861624911ccbe5361ce8bf6f90cae

75. Morgengryet  [Internet]. 1892 Mar 15:2. Available from: https://www.nb.no/items/334b2d6eb0f68ef98b03c773abfb1f6e

82. Fremskridt  [Internet]. 1892 Jan 15:2. Available from: https://www.nb.no/items/6e5fe9a803d9108c84c87326cac4a39c

83. Morsø Folkeblad (1877-) [Internet]. 1892 Jan 14:2. Available from: http://hdl.handle.net/109.3.1/uuid:6ee34aba-2ccd-4a93-9a92-2ed2bc29719f

84. Ærø Avis (1891-1963) [Internet]. 1892 Jan 14:2. Available from: http://hdl.handle.net/109.3.1/uuid:71f9647c-0c27-4b56-a163-bccc6f2a0874

85. Ribe Stifts-Tidende (1849-1960) [Internet]. 1892 Jan 15:2. Available from: http://hdl.handle.net/109.3.1/uuid:733e3d98-6e0d-4bca-9a01-e51877b84efa

86. Aalborg Stiftstidende (1885-1901) [Internet]. 1892 Jan 09:3. Available from: http://hdl.handle.net/109.3.1/uuid:7acafcf8-038e-4d18-ad13-d839dbb45b03

87. Holstebro Avis (1880-1895) [Internet]. 1892 Jan 13:3. Available from: http://hdl.handle.net/109.3.1/uuid:7c3eeb9c-e888-4e7a-ab74-a6645d126651

88. Skive Folkeblad (1880-) [Internet]. 1892 Jan 11:3. Available from: http://hdl.handle.net/109.3.1/uuid:7de9f482-1fc5-46ff-b382-f8d52f09665c

89. Horsens Arbejderblad (1888-1896) [Internet]. 1892 Jan 10:3. Available from: http://hdl.handle.net/109.3.1/uuid:7df4d75b-7696-4298-baaa-b136308c7855

90. Fyens Stiftstidende (1852-1993) [Internet]. 1892 Jan 12:2. Available from: http://hdl.handle.net/109.3.1/uuid:85ddb244-0365-4bc1-9770-117ec9a209e9

91. Aarhuus Stifts-Tidende (1871-1989) [Internet]. 1892 Jan 11:1. Available from: http://hdl.handle.net/109.3.1/uuid:86997db6-da2a-4d6c-b716-0db983ca455a

92. Horsens Folkeblad (1866-) [Internet]. 1892 Jan 11:2. Available from: http://hdl.handle.net/109.3.1/uuid:8c8797d2-026d-4896-96b6-8255527141ec

93. Stubbekøbing Avis (1867-1971) [Internet]. 1892 Jan 13:2. Available from: http://hdl.handle.net/109.3.1/uuid:8d50c935-24c5-4bc8-b5b7-8b5e3272762d

94. Tunsbergeren [Internet]. 1892 Mar 12:2. Available from: https://www.nb.no/items/9450b1bf63f6fb450c3b1c2122693071

101. Fredericia Dagblad (1890-) [Internet]. 1892 Jan 13:2. Available from: http://hdl.handle.net/109.3.1/uuid:dce4f4c0-245a-41fb-a4d7-3ee944ca1e60

102. Fredrikstad Dagblad [Internet]. 1892 Mar 07:2. Available from: https://www.nb.no/items/f88efd9264d7a8edb77006555d136b67

113. Morsø Folkeblad (1877-) [Internet]. 1893 Jan 05:2. Available from: http://hdl.handle.net/109.3.1/uuid:6a7cba8e-bc3e-4d55-8a1f-bfd603de7a85

114. Næstved Tidende. Sydsjællands Folkeblad (1866-1918) [Internet]. 1893 Jan 05:2. Available from: http://hdl.handle.net/109.3.1/uuid:6b25e401-bcab-4963-afec-b66cc84bade5

115. Kjøge Avis (1874-1899) [Internet]. 1893 Jan 06:2. Available from: https://www2.statsbiblioteket.dk/mediestream/avis/record/doms_aviser_page:uuid:74465803-4d21-42f2-8f56-8786b554754f/query/%22tungen%20fast%22~3

116. Slagelse-Posten (1867-1916) [Internet]. 1893 Jan 05:2. Available from: http://hdl.handle.net/109.3.1/uuid:754aecac-6664-497b-8ebd-303c66495497

117. Frederiksborg Amts Avis (1874-) [Internet]. 1893 Jan 08:2. Available from: https://www2.statsbiblioteket.dk/mediestream/avis/record/doms_aviser_page:uuid:7629ed0c-070b-493b-9e05-37ffeee04d50/query/%22tunge%20fast%22~3

124. Fredericia Dagblad (1890-) [Internet]. 1893 Jan 04:2. Available from: http://hdl.handle.net/109.3.1/uuid:cb07b148-8c87-40a5-b1b4-bf6452cc5c0d

125. Korsør Avis (1855-1957) [Internet]. 1893 Jan 06:2. Available from: https://www2.statsbiblioteket.dk/mediestream/avis/record/doms_aviser_page:uuid:dabdfc3d-747e-4665-98f6-b5e6ed636997/query/%22tunge%20fast%22~3

126. Stubbekøbing Avis (1867-1971) [Internet]. 1893 Jan 06:2. Available from: http://hdl.handle.net/109.3.1/uuid:dca442f1-5589-4510-8e9d-d193d05dbc98

127. Svendborg Avis. Sydfyns Tidende (1869-1970) [Internet]. 1893 Jan 06:2. Available from: http://hdl.handle.net/109.3.1/uuid:e3ca0819-9eb0-4882-8d20-f6b345f3ac4f

128. Vejle Amts Folkeblad (1865-) [Internet]. 1893 Jan 04:2. Available from: https://www2.statsbiblioteket.dk/mediestream/avis/record/doms_aviser_page:uuid:e679793f-f414-417f-b830-a4aaacc948d5/query/%22tunge%20fast%22~3

129. Randers Dagblad og Folketidende (1874-1970) [Internet]. 1893 Jan 07:2. Available from: http://hdl.handle.net/109.3.1/uuid:e7550716-e7d8-40cf-b798-ca418369f755

130. Randers Amtsavis og Adressecontoirs Efterretninger (1857-1947) [Internet]. 1893 Jan 05:2. Available from: https://www2.statsbiblioteket.dk/mediestream/avis/record/doms_aviser_page:uuid:e7c0c329-eb63-4ab0-9749-a932951d5237/query/%22tunge%20fast%22~3

131. Horsens Folkeblad (1866-) [Internet]. 1893 Jan 05:2. Available from: https://www2.statsbiblioteket.dk/mediestream/avis/record/doms_aviser_page:uuid:fab6b15c-6b09-4abd-afc7-92b948f06361/query/%22tunge%20fast%22~3

132. Reformatorn [Internet]. 1895 Feb 14:3. Available from: https://tidningar.kb.se/1kcqvmvc0txw1j0/part/1/page/3

133. Vårt land Weckupplaga [Internet]. 1895 Feb 14:1. Available from: https://tidningar.kb.se/2jtbph1h084cpcfm/part/1/page/1

134. Hvetlanda tidning [Internet]. 1895 Feb 11:3. Available from: https://tidningar.kb.se/3klf4qvp1lpq2xjj/part/1/page/3

135. Fäderneslandet (Stockholm : 1852) [Internet]. 1895 Feb 09:3. Available from: https://tidningar.kb.se/4g80m3qc2hjkz02g/part/1/page/3

136. Folkets tidning [Internet]. 1895 Feb 11:3. Available from: https://tidningar.kb.se/5jh1dwvx30vr9pv2/part/1/page/3

137. Oscarshamnsposten [Internet]. 1895 Feb 09:3. Available from: https://tidningar.kb.se/6n0bng6m48xsdgjq/part/1/page/3

138. Öresundsposten (Ängelholm : 1847) [Internet]. 1895 Feb 13:2. Available from: https://tidningar.kb.se/7lwphn8x5vz2537f/part/1/page/2

139. Norrbottens allehanda [Internet]. 1895 Feb 15:3. Available from: https://tidningar.kb.se/7p86s7nc5qj67wrv/part/1/page/3

140. Fosterlandet (1886) [Internet]. 1895 Feb 14:2. Available from: https://tidningar.kb.se/8m9sqhnw6mbbg8pj/part/1/page/2

141. Tidning för Wenersborgs stad och län [Internet]. 1895 Feb 08:3. Available from: https://tidningar.kb.se/g0s52n3s2vkxl2x/part/1/page/3

142. Nya Norrlänningen [Internet]. 1895 Feb 16:4. Available from: https://tidningar.kb.se/hs412tzbf6xx40ck/part/1/page/4

143. Mariefreds nya tidning [Internet]. 1895 Feb 08:2. Available from: https://tidningar.kb.se/js8zw6txg6948nwk/part/1/page/2

144. Södertälje tidning [Internet]. 1895 Feb 08:3. Available from: https://tidningar.kb.se/kzksn28qhnljktlp/part/1/page/3

145. Eslöfs tidning [Internet]. 1895 Feb 09:2. Available from: https://tidningar.kb.se/p1vhgv3smdd6d2ht/part/1/page/2

146. Köpingsposten [Internet]. 1895 Feb 13:3. Available from: https://tidningar.kb.se/p2jrf86lmvxzbvbn/part/1/page/3

147. Södermanlands läns tidning [Internet]. 1895 Feb 07:3. Available from: https://tidningar.kb.se/p4k56k79mjszf829/part/1/page/3

148. Helsingen (Söderhamn : 1882) [Internet]. 1895 Feb 09:3. Available from: https://tidningar.kb.se/q1fq29plnv13vfzf/part/1/page/3

149. Söderköpingsposten [Internet]. 1895 Feb 08:3. Available from: https://tidningar.kb.se/q5zcwjdwnfg6xh4p/part/1/page/3

150. Östergötlands-Södermanlands annonsblad [Internet]. 1895 Feb 09:2. Available from: https://tidningar.kb.se/s2bqqq9lqqcfd2w9/part/1/page/2

151. Södermanlands nyheter [Internet]. 1895 Feb 06:3. Available from: https://tidningar.kb.se/s8whfvfnq46hcrlr/part/1/page/3

152. Helsingborgsposten Skåne Halland [Internet]. 1895 Feb 09:2. Available from: https://tidningar.kb.se/wct2455vtgwk27vc/part/1/page/2

153. Bergslagsposten (Lindesberg : 1892-) [Internet]. 1895 Feb 09:3. Available from: https://tidningar.kb.se/x6h9047gv4pkprz9/part/1/page/3

154. Söderhamns tidning [Internet]. 1895 Feb 11:3. Available from: https://tidningar.kb.se/xg8pqft83hf6szw/part/1/page/3

155. Östergötlands dagblad [Internet]. 1895 Feb 06:2. Available from: https://tidningar.kb.se/zd8mwz9hw8dj10tg/part/1/page/2

156. Karlshamn [Internet]. 1895 Feb 08:3. Available from: https://tidningar.kb.se/zdth0xw3wk6573f2/part/1/page/3

157. Randers Amtsavis og Adressecontoirs Efterretninger (1857-1947) [Internet]. 1895 Feb 05:2. Available from: http://hdl.handle.net/109.3.1/uuid:1928dbd5-3bea-4d32-8b88-58bdb473e667

158. Fyens Stiftstidende (1852-1993) [Internet]. 1895 Feb 04:2. Available from: http://hdl.handle.net/109.3.1/uuid:27c2b887-e17d-4e17-9c2d-6caf3b149025

159. Ærø Avis (1891-1963) [Internet]. 1895 Feb 05:2. Available from: http://hdl.handle.net/109.3.1/uuid:2aafebfb-e808-4103-a985-0357b436b332

160. Skånes annonsblad [Internet]. 1895 Feb 09:3. Available from: https://tidningar.kb.se/3hp5jmp81xwl89nj/part/1/page/3

161. Viborg Stifts-Tidende (1876-1962) [Internet]. 1895 Feb 05:2. Available from: http://hdl.handle.net/109.3.1/uuid:41eddedd-cd2c-47ce-af3b-2680225f6e0e

162. Thisted Amtsavis (1859-1960) [Internet]. 1895 Feb 02:2. Available from: http://hdl.handle.net/109.3.1/uuid:7205eaf9-9850-4cc7-bd85-6bd7191977d2

163. Helsingborgsposten Skåne Halland [Internet]. 1895 Feb 07:3. Available from: https://tidningar.kb.se/8q6fhhdx6h189nk9/part/1/page/3

164. Kongelig allernaadigst privilegeret Horsens Avis eller Skanderborg Amtstidende (1842-1961) [Internet]. 1895 Feb 04:2. Available from: http://hdl.handle.net/109.3.1/uuid:a27907f5-8971-4ec4-8a73-5a13ff33550e

165. Ribe Stifts-Tidende (1849-1960) [Internet]. 1895 Feb 05:1. Available from: http://hdl.handle.net/109.3.1/uuid:dd2fd749-4814-4cbc-ba69-7fed8de316e4

166. Vejle Amts Folkeblad (1865-) [Internet]. 1895 Feb 02:2. Available from: http://hdl.handle.net/109.3.1/uuid:de6eb192-8b97-4a9e-9e15-c48ff14f4a19

167. Aarhus Amtstidende (1866-1965) [Internet]. 1895 Jan 30:2. Available from: http://hdl.handle.net/109.3.1/uuid:e3c40b52-7349-40a7-a821-0d10d0e2425f

168. Borås tidning [Internet]. 1895 Feb 09:3. Available from: https://tidningar.kb.se/sb4gwt541njv91v/part/1/page/3

169. Ölandsbladet [Internet]. 1895 Mar 02:3. Available from: https://tidningar.kb.se/s406g063qwdx7ksf/part/1/page/3

170. Socialdemokraten [Internet]. 1895 Feb 21:3. Available from: https://tidningar.kb.se/08cj1pmnxm1v5lpt/part/1/page/3

171. Ny tid [Internet]. 1895 Mar 01:4. Available from: https://tidningar.kb.se/0bkj5c43x7m5tnlv/part/1/page/4

172. Göteborgs aftonblad (1888) [Internet]. 1895 Feb 20:3. Available from: https://tidningar.kb.se/2ldnqqzd2jpt8p3/part/1/page/3

173. Skåningen Eslövs tidning [Internet]. 1895 Feb 28:4. Available from: https://tidningar.kb.se/8lm1hmsh60k9604h/part/1/page/4

174. Dannebrog (København) (1892-1910) [Internet]. 1895 Feb 15:3. Available from: http://hdl.handle.net/109.3.1/uuid:c20df8d2-180f-485a-add0-1e057a949585

175. Smålands allehanda [Internet]. 1895 Feb 18:2. Available from: https://tidningar.kb.se/p4bdjtb7mzlqjp2k/part/1/page/2

176. Ystads allehanda [Internet]. 1895 Feb 23:3. Available from: https://tidningar.kb.se/s8h224fnqxbmnxx0/part/1/page/3

177. Gotlänningen [Internet]. 1895 Mar 02:2. Available from: https://tidningar.kb.se/v9gmlpsmsf5znknz/part/1/page/2

178. Vestlandske Tidende [Internet]. 1895 Feb 13:. Available from: https://www.nb.no/items/2e8daa8f095bb498eed7d31bdac8930d

179. Kristianiaposten [Internet]. 1895 Feb 09:. Available from: https://www.nb.no/items/5f9f8a9eff1d6ee45c13644a7499b0bc

180. Kongsberg Blad [Internet]. 1895 Feb 08:. Available from: https://www.nb.no/items/7e72b5863efcb7423cebd13d0a12fccf

181. Bergens Annonce Tidende [Internet]. 1895 Feb 11:. Available from: https://www.nb.no/items/ced2186db4496626d0a99987a4a6f133

182. Stockholms nyheter [Internet]. 1895 Feb 16:3. Available from: https://tidningar.kb.se/0gpmf450xrxr727x/part/1/page/3

183. Engelholms tidning (1867) [Internet]. 1895 Feb 16:3. Available from: https://tidningar.kb.se/3gd2dlzv1xxt6pm4/part/1/page/3

184. Helsingborgsposten Skåne Halland [Internet]. 1895 Feb 13:2. Available from: https://tidningar.kb.se/7p5dgf0954nz650k/part/1/page/2

185. Skånska dagbladet [Internet]. 1895 Feb 12:2. Available from: https://tidningar.kb.se/brwkj3cf8bpn3w8s/part/1/page/2

186. Upsala [Internet]. 1895 Feb 14:3. Available from: https://tidningar.kb.se/dxq2kkcq3tksvj0/part/1/page/3

187. Søndenfjeldske Avis [Internet]. 1897 Feb 20:1. Available from: https://www.nb.no/items/616874c36b448e6b7e9808ab5a1aebc7

188. Farsunds Avis [Internet]. 1897 Feb 23:2. Available from: https://www.nb.no/items/860558556cd251a7850d18e3a2382e6c

189. Lister og Mandals Amtstidende og Adresseavis [Internet]. 1897 Feb 25:2. Available from: https://www.nb.no/items/88e71d4f48dccc3f022f8f43115d8531

190. Lindesnes [Internet]. 1897 Feb 17:. Available from: https://www.nb.no/items/a3d416bea2a822d17bbf2af0fce1ef2b

191. Dagsposten [Internet]. 1897 Feb 09:4. Available from: https://www.nb.no/items/d99645ba19c66b71759914f8cae6df8f

192. Blekingekuriren (Karlskrona : 1892) [Internet]. 1898 Feb 02:3. Available from: https://tidningar.kb.se/0dzqlqdgxxg481ds/part/1/page/3

193. Skånska dagbladet [Internet]. 1898 Feb 02:3. Available from: https://tidningar.kb.se/0fdldmklxb6tshdg/part/1/page/3

194. Smålands allehanda [Internet]. 1898 Feb 03:3. Available from: https://tidningar.kb.se/0fmlh368xjp5jq1x/part/1/page/3

195. Skånska posten [Internet]. 1898 Feb 02:4. Available from: https://tidningar.kb.se/1dh3fvk0zvlwbprs/part/1/page/4

196. Nerikes allehanda [Internet]. 1898 Feb 02:3. Available from: https://tidningar.kb.se/1dt5kndxz55sdshz/part/1/page/3

197. Falukuriren [Internet]. 1898 Feb 08:3. Available from: https://tidningar.kb.se/2bg1m5bv0wgh3wfw/part/1/page/3

198. Norrköpings tidningar [Internet]. 1898 Feb 01:3. Available from: https://tidningar.kb.se/2cr4nw7k0j5pczwm/part/1/page/3

199. Jämtlands tidning (Östersund : 1895) [Internet]. 1898 Feb 02:3. Available from: https://tidningar.kb.se/2ffj2bh205z04k1r/part/1/page/3

200. Umeå nya tidning [Internet]. 1898 Feb 04:3. Available from: https://tidningar.kb.se/2gqm48p20dlt0nns/part/1/page/3

201. Norrlandsposten (1837) [Internet]. 1898 Jan 31:3. Available from: https://tidningar.kb.se/3cdkpwt519242fhr/part/1/page/3

202. Helsingborgs dagblad [Internet]. 1898 Feb 03:2. Available from: https://tidningar.kb.se/3d0s0ck71l88kj6l/part/1/page/2

203. Förposten [Internet]. 1898 Feb 05:1. Available from: https://tidningar.kb.se/4ktj070d2hxftf4c/part/1/page/1

204. Ystads allehanda [Internet]. 1898 Feb 05:3. Available from: https://tidningar.kb.se/5mvfn7343hvwttmw/part/1/page/3

205. Skara tidning [Internet]. 1898 Feb 05:3. Available from: https://tidningar.kb.se/5mvpk2803ntdmkb9/part/1/page/3

206. Sörmlandsposten [Internet]. 1898 Jan 31:4. Available from: https://tidningar.kb.se/6n9sh05w4dh33js3/part/1/page/4

207. Boråsposten veckoupplagan (1894) [Internet]. 1898 Feb 05:3. Available from: https://tidningar.kb.se/6nfxnk8w4tkmx4xm/part/1/page/3

208. Eskilstunakuriren [Internet]. 1898 Feb 04:3. Available from: https://tidningar.kb.se/6qjw2vnj28kb3xz/part/1/page/3

209. Hudiksvallsposten [Internet]. 1898 Feb 01:3. Available from: https://tidningar.kb.se/7k4q4f4x5tvcbqzf/part/1/page/3

210. Arvika tidning [Internet]. 1898 Feb 04:3. Available from: https://tidningar.kb.se/7m2n7c2j55xkjzsv/part/1/page/3

211. Södermanlands läns tidning [Internet]. 1898 Jan 31:3. Available from: https://tidningar.kb.se/7n3t0w3t5v6sgpct/part/1/page/3

212. Skara annonsblad (Skara : 1891) [Internet]. 1898 Feb 05:3. Available from: https://tidningar.kb.se/9n11252z7qz40xwz/part/1/page/3

213. Vårt land (Stockholm : 1886) [Internet]. 1898 Jan 29:3. Available from: https://tidningar.kb.se/9qk9l252713jl7mn/part/1/page/3

214. Hessleholms tidning (Kristianstad : 1889) [Internet]. 1898 Feb 01:3. Available from: https://tidningar.kb.se/bnnhl4wx8v1mcfnq/part/1/page/3

215. Nya Dagligt Allehanda [Internet]. 1898 Jan 31:4. Available from: https://tidningar.kb.se/bscfw9dv8dz0bhjz/part/1/page/4

216. Östgötakuriren (Vadstena : 1883) [Internet]. 1898 Feb 05:3. Available from: https://tidningar.kb.se/cq8h95ss9x57r95d/part/1/page/3

217. Enköpings tidning (1885) [Internet]. 1898 Feb 02:3. Available from: https://tidningar.kb.se/cs2nnr4p9gms5vsd/part/1/page/3

218. Karlskoga tidning [Internet]. 1898 Feb 04:2. Available from: https://tidningar.kb.se/dp5rfnw4b1w60trg/part/1/page/2

219. Karlstadstidningen [Internet]. 1898 Feb 02:3. Available from: https://tidningar.kb.se/dr19ncpxbsdzztpx/part/1/page/3

220. Avestaposten [Internet]. 1898 Feb 01:4. Available from: https://tidningar.kb.se/dst6xbb9b3lzlmm3/part/1/page/4

221. Härnösandsposten [Internet]. 1898 Jan 27:2. Available from: https://tidningar.kb.se/fvmv3cx5cw8qk6pj/part/1/page/2

222. Lund [Internet]. 1898 Feb 02:2. Available from: https://tidningar.kb.se/gt4vnb5gdfk55qt4/part/1/page/2

223. Skånetidningen [Internet]. 1898 Feb 04:3. Available from: https://tidningar.kb.se/gt80znq0d3f38jtx/part/1/page/3

224. Boråsposten [Internet]. 1898 Feb 04:3. Available from: https://tidningar.kb.se/ht66rhs0fv26mb3l/part/1/page/3

225. Wadstena läns tidning [Internet]. 1898 Feb 03:3. Available from: https://tidningar.kb.se/jzclsvxjg99nlwcx/part/1/page/3

226. Upsala [Internet]. 1898 Feb 01:3. Available from: https://tidningar.kb.se/jzs1brrng8zg3mtv/part/1/page/3

227. Söderhamnskuriren (1895) [Internet]. 1898 Feb 02:3. Available from: https://tidningar.kb.se/jzv71z8pgtw3xdvb/part/1/page/3

228. Blekinge läns tidning [Internet]. 1898 Feb 02:2. Available from: https://tidningar.kb.se/kw8hbt3hhfd33r84/part/1/page/2

229. Nya Kristinehamnsposten [Internet]. 1898 Feb 04:4. Available from: https://tidningar.kb.se/kzh4wgs1h37f9c8l/part/1/page/4

230. Bärgslagsbladet (Sala : 1890) [Internet]. 1898 Feb 04:3. Available from: https://tidningar.kb.se/l0d99mncjdfxdchf/part/1/page/3

231. Östra Westmanland [Internet]. 1898 Feb 01:3. Available from: https://tidningar.kb.se/l1zqj9kljwlshlmz/part/1/page/3

232. Nora stads och Bergslags tidning [Internet]. 1898 Feb 02:3. Available from: https://tidningar.kb.se/lzdrrk1vjkggzl3h/part/1/page/3

233. Mariestads länstidning [Internet]. 1898 Feb 04:3. Available from: https://tidningar.kb.se/m152kxx7kp5vr8kk/part/1/page/3

234. Skärgården (Uddevalla : 1897) [Internet]. 1898 Feb 02:2. Available from: https://tidningar.kb.se/m3k77pddklx5pl8l/part/1/page/2

235. Karlstadstidningen Veckoupplagan [Internet]. 1898 Feb 02:3. Available from: https://tidningar.kb.se/m3kpxgxzk1cmqh54/part/1/page/3

236. Skåningen Eslövs tidning [Internet]. 1898 Feb 03:2. Available from: https://tidningar.kb.se/n0wwtngql5xt26n1/part/1/page/2

237. Nya Stockholmsposten [Internet]. 1898 Feb 04:1. Available from: https://tidningar.kb.se/n13qpp8ml4hjqbzq/part/1/page/1

238. Östgöta correspondenten [Internet]. 1898 Jan 31:3. Available from: https://tidningar.kb.se/n3zf52l5l1f19jd0/part/1/page/3

239. Engelholms tidning (1867) [Internet]. 1898 Feb 03:2. Available from: https://tidningar.kb.se/p2zjk6w1mt3gxdck/part/1/page/2

240. Hörbyposten centralskåne [Internet]. 1898 Feb 03:2. Available from: https://tidningar.kb.se/q3vr2xkzn3p0lngt/part/1/page/2

241. Dagen (Stockholm : 1896) [Internet]. 1898 Jan 31:3. Available from: https://tidningar.kb.se/r3q4jd3cp3b7x7wq/part/1/page/3

242. Borås tidning [Internet]. 1898 Feb 01:2. Available from: https://tidningar.kb.se/r3sq7cbgpj0bn6lx/part/1/page/2

243. Veckoposten [Internet]. 1898 Feb 03:3. Available from: https://tidningar.kb.se/r60d291spghdlqh5/part/1/page/3

244. Skaraborgs läns tidning (Skövde : 1884) [Internet]. 1898 Feb 05:3. Available from: https://tidningar.kb.se/r60th4mrpnn54xl7/part/1/page/3

245. Jämtlandsposten [Internet]. 1898 Jan 31:2. Available from: https://tidningar.kb.se/r93jfdk302jgz4t/part/1/page/2

246. Socialdemokraten [Internet]. 1898 Feb 01:3. Available from: https://tidningar.kb.se/s26570vjq7fzr2cs/part/1/page/3

247. Tranåsposten (Linköping : 1898) [Internet]. 1898 Feb 04:3. Available from: https://tidningar.kb.se/s4x279qfq21dlqv0/part/1/page/3

248. Vestgöten (Borås : 1888) [Internet]. 1898 Feb 04:3. Available from: https://tidningar.kb.se/s6ghq71hq7khvtcg/part/1/page/3

249. Elfsborgs läns annonsblad [Internet]. 1898 Feb 08:3. Available from: https://tidningar.kb.se/s7g43x4wqx1vhs42/part/1/page/3

250. Nya Norrlänningen [Internet]. 1898 Feb 02:2. Available from: https://tidningar.kb.se/t4blj47brc75wldk/part/1/page/2

251. Falkenbergs tidning [Internet]. 1898 Feb 09:2. Available from: https://tidningar.kb.se/t9l4j5h5r84f7763/part/1/page/2

252. Nya Wexjöbladet [Internet]. 1898 Feb 01:3. Available from: https://tidningar.kb.se/t9wmw8kfr25r2rr1/part/1/page/3

253. Folkets tidning [Internet]. 1898 Feb 02:3. Available from: https://tidningar.kb.se/v76q5z9cs4877hgf/part/1/page/3

254. Sölvesborgsposten [Internet]. 1898 Feb 02:2. Available from: https://tidningar.kb.se/v8j30hczs5kfb7d5/part/1/page/2

255. Grythytte tidning [Internet]. 1898 Feb 04:2. Available from: https://tidningar.kb.se/vb3jwvh7sdr0mjbt/part/1/page/2

256. Dagens nyheter [Internet]. 1898 Jan 31:1. Available from: https://tidningar.kb.se/vd6c66962vb51kr/part/1/page/1

257. Smålandsbladet (Eksjö : 1898) [Internet]. 1898 Feb 04:3. Available from: https://tidningar.kb.se/x9mbfs6cvtgnxmdn/part/1/page/3

258. Cimbrishamnsbladet [Internet]. 1898 Feb 09:2. Available from: https://tidningar.kb.se/xcjvj2cjvqprtkc4/part/1/page/2

259. Helsingborgsposten Skåne Halland [Internet]. 1898 Jan 31:2. Available from: https://tidningar.kb.se/xdvxl9fqv0lmqqv6/part/1/page/2

260. Motala tidning (1868) [Internet]. 1898 Feb 02:2. Available from: https://tidningar.kb.se/xdwr9mkqv0t29dsl/part/1/page/2

261. Kristinehamnstidningen [Internet]. 1898 Feb 03:3. Available from: https://tidningar.kb.se/z7n08dhwwb0c6l61/part/1/page/3

262. Skelleftebladet [Internet]. 1898 Feb 07:3. Available from: https://tidningar.kb.se/zd87ts43wpg1t3l6/part/1/page/3

263. Åmålsposten [Internet]. 1898 Feb 05:3. Available from: https://tidningar.kb.se/zfw14dpzwqj8b4qd/part/1/page/3

264. Upsala [Internet]. 1898 Nov 26:3. Available from: https://tidningar.kb.se/3jclzl931tjxk3w0/part/1/page/3

265. Ljusdals tidning [Internet]. 1898 Dec 01:4. Available from: https://tidningar.kb.se/5jvhdsq63t6k42sn/part/1/page/4

266. Hudiksvallsposten [Internet]. 1898 Nov 29:3. Available from: https://tidningar.kb.se/7k4qp61x5ccn72ng/part/1/page/3

267. Södermanlands läns tidning [Internet]. 1898 Nov 28:3. Available from: https://tidningar.kb.se/br6t9gdv867rhs2l/part/1/page/3

268. Tidning för Falu län och stad [Internet]. 1898 Nov 30:3. Available from: https://tidningar.kb.se/drfn5j6qb38d9vs7/part/1/page/3

269. Gotlands allehanda [Internet]. 1898 Nov 28:2. Available from: https://tidningar.kb.se/jtkqmdt4g8d0s5hh/part/1/page/2

270. Bergslagsposten (Lindesberg : 1892-) [Internet]. 1898 Dec 03:3. Available from: https://tidningar.kb.se/mw4jlgmhkjb4wv82/part/1/page/3

271. Gotland [Internet]. 1898 Nov 28:2. Available from: https://tidningar.kb.se/zf7msfskw9rch533/part/1/page/2

272. Östgöten (Linköping : 1874) [Internet]. 1899 Dec 27:3. Available from: https://tidningar.kb.se/r37z0sbrppbqx76z/part/1/page/3
